# Three-dimensional topography of Descemet’s membrane in Fuchs endothelial corneal dystrophy using laser scanning confocal microscopy and white-light interferometry

**DOI:** 10.64898/2026.04.07.26350293

**Authors:** Corantin Maurin, Sylvain Poinard, Gauthier Travers, Zhiguo He, Fanny Decoeur, Etienne Gontier, Georgia Karpathiou, Philippe Gain, Gilles Thuret, the French Fuchs Study Group (FFSG)

## Abstract

**Aim:** To evaluate the potential of a three-dimensional microscope combining Laser scanning confocal imaging and white-light interferometry for quantitative topographic characterisation of Descemet’s membrane (DM) in Fuchs endothelial corneal dystrophy (FECD).

**Methods:** Descemet’s membranes were collected from 38 FECD patients undergoing endothelial keratoplasty and 4 healthy donors. After flat-mounting on glass slide and drying, specimens were analysed using the VK-X3000 system (KEYENCE). Entire samples were reconstructed by image stitching at low magnification (×10) in white-light interferometry mode (0.01nm axial resolution). Higher magnifications (×20-×150) in confocal mode (12 nm axial resolution) enabled detailed structural analysis. Three-dimensional height maps were generated to calculate standardised surface roughness parameters. Guttae and other DM features were classified according to spatial organisation and elevation profiles.

**Results:** White-light interferometry enabled full-field mapping of whole 8mm diameter DMs with nanometric vertical resolution (∼2 hours/sample). Surface roughness (Sa) was higher in FECD than in controls (median±IQR: 0.571±0.259 µm vs 0.239±0.161 µm; p = 0.0018). In FECD, three zones were identified: central (guttae buried in the posterior fibrillar layer; Sa 0.442 ± 0.112 µm), paracentral (large uncovered guttae; Sa 0.562±0.170 µm; p = 0.0423), and outer zone (no confluent guttae; Sa 0.261±0.143 µm; p < 0.0001). Confocal 3D imaging revealed radial striae, embossments and furrows in the DM, confluent central guttae, and fused or buried structures.

**Conclusions:** Combining white-light interferometry and confocal microscopy enables label-free, high-resolution surface characterisation of DM in FECD, providing quantitative metrics to compare histological subtypes and supporting the predominance of radial structural organisation.

## INTRODUCTION

Fuchs endothelial corneal dystrophy (FECD) is the leading indication for corneal transplantation worldwide [1]. It affects at diverse stages 4% to 12% of adults in Western countries [2]. Its prevalence is increasing due to population ageing [3], and it has been estimated that 10% to 20% of affected individuals may ultimately require corneal transplantation [4]. FECD is characterised by progressive dysfunction and premature loss of corneal endothelial cells (CECs), which in some patients leads -after decades of evolution-to permanent corneal oedema that is reversible after transplantation.

In addition to endothelial failure, earlier visual symptoms are partly driven by alterations of Descemet’s membrane (DM), the CEC basement membrane [2], which modify corneal optical properties. These changes are of two main types: (i) diffuse extracellular matrix (ECM) accumulation leading to a global increase in DM thickness, and (ii) focal deposits known as guttae. The mechanisms and dynamics of ECM remodelling remain incompletely understood. The dramatic transformation of a normally smooth membrane likely imposes new mechanobiological constraints on CECs and contributes to a vicious cycle in which cellular and matrix abnormalities mutually reinforce each other [5].

Several aspects of guttae have been described in the literature, including size, shape, arrangement, location, and association with other DM changes [6]-[7]. This diversity may reflect not only differences in analytical methods but also the genuine heterogeneity of guttae, both within the same DM as well as between patients [8]. However, available data remain limited by the technical constraints of imaging methods employed. Electron microscopy, although highly detailed, requires lengthy sample preparation [9] and only allows observation of a restricted area, most often in cross-section [10]. Conversely, non-invasive techniques such as specular microscopy [11] [12] or confocal microscopy [13] provide *in vivo* visualisation but lack three-dimensional resolution to fully capture the complexity of guttae and their distribution within DM. This lack of imaging tools combining high resolution with three-dimensional assessment over large areas hinders detailed understanding of Descemetic changes and their exact role in FECD progression.

The VK-X3000 microscope (Keyence, Osaka, Japan) is a 3D optical profilometer originally used in the electronics and material sciences but increasingly applied in biological studies to characterise surface topography [14],[15]. It integrates two particularly relevant optical modalities for DM surface characterisation: 1/ Laser scanning confocal microscopy, which measures surface topography based on the reflected laser signal, across a range of magnifications depending on the objective (×4 to ×150) 2/ White-light interferometry, which operates by analysing interference between a reference arm and an arm interacting with the specimen, measures very fine height variations (sub-nanometre axial sensitivity) at low magnification (×10), thereby enabling wide-field acquisition. In both imaging modes, the instrument generates topographic maps allowing height and roughness parameter quantification as defined by international standards[16]. In addition, an image-stitching mode allows adjacent tiles to be assembled to cover large areas, from a few micrometres up to 50 mm × 50 mm depending on the imaging modality.

In the present study, we leveraged this microscope to analyse DM surface topography in FECD. Our objective was to demonstrate the potential of these measurements for characterising elementary FECD lesions and for classifying distinct histological phenotypes.

## MATERIALS AND METHODS

### Sample preparation

Pathological specimens were sourced from the FECD 500 reference collection through the French Fuchs Study Group (FFSG, Supplementary 1). These cases, recruited from various FFSG-affiliated ophthalmology centres, represent the diverse clinical and pathological phenotypes of FECD which we have recently characterised and classified [8]. A total of 38 patients with FECD were included (mean age 68 ± 8 (Standard deviation) years; male/female ratio 0.35); of these, 34 complete DM were used for statistical analysis, while the remaining 4 samples, as they were incomplete, were used to image specific structures of interest at high confocal magnification. Samples (approximately 8 mm in diameter) was obtained by Descemetorhexis, a surgical technique consisting in removing the central pathological endothelium at the beginning of corneal graft. All specimens used from this collection were detailed in the Supplementary Material (Supplementary 2).

Healthy corneas were also analysed, obtained from 4 donors from the anatomy laboratory of the Faculty of Medicine of Saint-Étienne (body donation to Science), with a mean age of 73 ± 14.5 years and a male /female ratio = 1. Preliminary light microscopy observation was performed to confirm the absence of endothelial diseases [17]. The endothelium was then peeled off using a standardised « no-touch » technique and trephined at 8 mm diameter [18].

Depending on the collection centre, samples were immersed in ultrapure water, phosphate-buffered saline (PBS), or 0.5% paraformaldehyde (PFA) and transported to the laboratory at room temperature (Supplementary 2). All samples in water and BSS were totally decellularized and the samples in PFA were largely deprived from their CECs allowing focusing solely on the matrix component of the disease. A single sample from a healthy donor was kept cellularised in order to prove the feasibility of the concept for future studies (Fig 1B).

**Figure 1.**
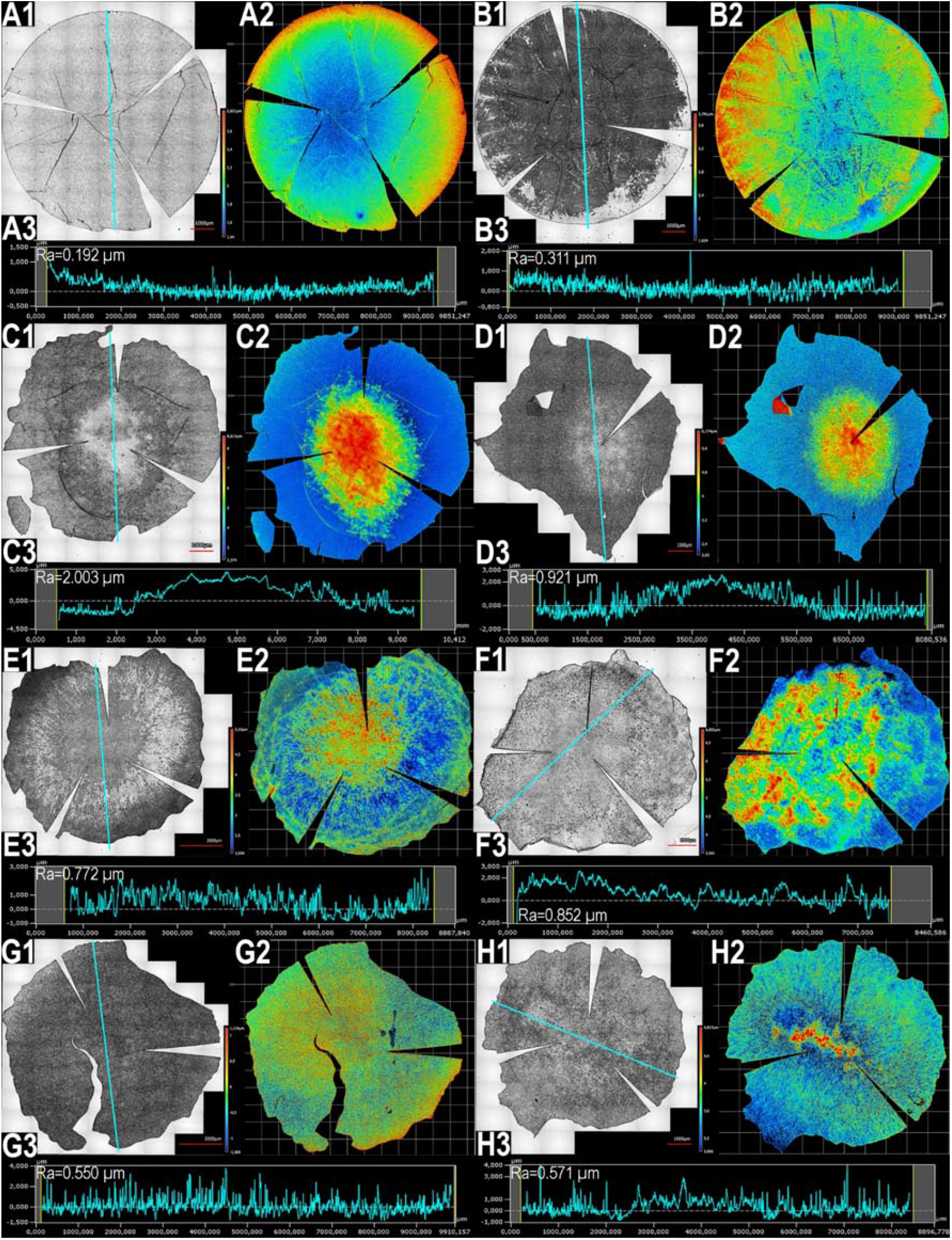
Nano-topography (×10 white-light interferometry) of whole Descemet’s membranes (DM) from healthy donors and FECD patients. For each specimen: **(1):** brightfield image of the DM (scale bar: 1000 µm in A, B, C, D, F, H; 2000 µm in E, G); **(2):** 3D elevation (height) map; **(3):** linear height profile used for roughness analysis (blue line in (1)). (Ra): linear arithmetic mean height of the profile. **(A):** Acellular DM from a healthy donor. **(B):** DM from a healthy donor with endothelial cells. **(C–G):** Representative decellularised FECD DMs showing heterogeneous guttae distribution.

Upon receipt, DMs were rinsed in PBS and flat-mounted on glass slides (Series 2 adhesive, Trajan Scientific, Ringwood, Australia) under an operating microscope, endothelial side up, using ultrapure water to avoid crystallisation artefacts during drying. Two to four radial incisions were made with a scalpel blade to allow flat mounting with minimal folds. Water was then absorbed using a micro-sponge placed at the edge of the DM to ensure complete adhesion to the slide. Flat-mounted tissue was air-dried at RT. Slides were stored at 4°C in an airtight container until 3D topography analysis.

### Microscope and image acquisition

Samples were imaged using a laser scanning confocal microscope (CLSM) coupled with white-light interferometry (VK-X3000 series, KEYENCE Corporation, Osaka, Japan). The system included the controller (VK-X3000), a 404-nm laser confocal head (VK-X3100), a stand with a motorised XY stage (100 × 100 mm, VK-D3), a goniometric stage (OP-88549), and a set of objectives (10x to 150x) providing different magnification (References and resolutions in Supplementary 3).

Image acquisition was performed with VK-X3000 Viewer (VK-A3E, version 2.3.0.151). Image stitching was performed using VK-X3000 Laser Image Stitching (VK-H3J, version 3.4.0) for CLSM images and VK-X3000 Image Stitching (VK-H3I, version 2.4.0.55) for white-light interferometry images. Post-processing and surface analyses were performed using VK-X3000 MultiFileAnalyzer (version 3.3.1.85).

### Image Post Processing and Surface Roughness parameters

Regardless of the optical modality, images could be generated in three formats: (1) Brightfield images corresponding to reconstructed reflected-intensity images from the scanned region of interest; (2) Height maps corresponding to topography images derived from laser scanning or withe-light interferometry, with the z position of each pixel determined from the maximum reflection signal; (3) C-DIC images, which are virtual transformations of brightfield images to approximate differential interference contrast (DIC).

A pre-processing step was performed to verify and, when necessary, correct the reference plane in cases of sample tilt relative to the glass slide (assumed to be planar). An additional processing step (“wavefront removal”) was applied to high-magnification images to emphasise local surface features by removing global topography trends (Supplementary 4) in particular to isolate the pattern of curly fibres (Fig. 4, 3D-flattened images).

**Figure 2.**
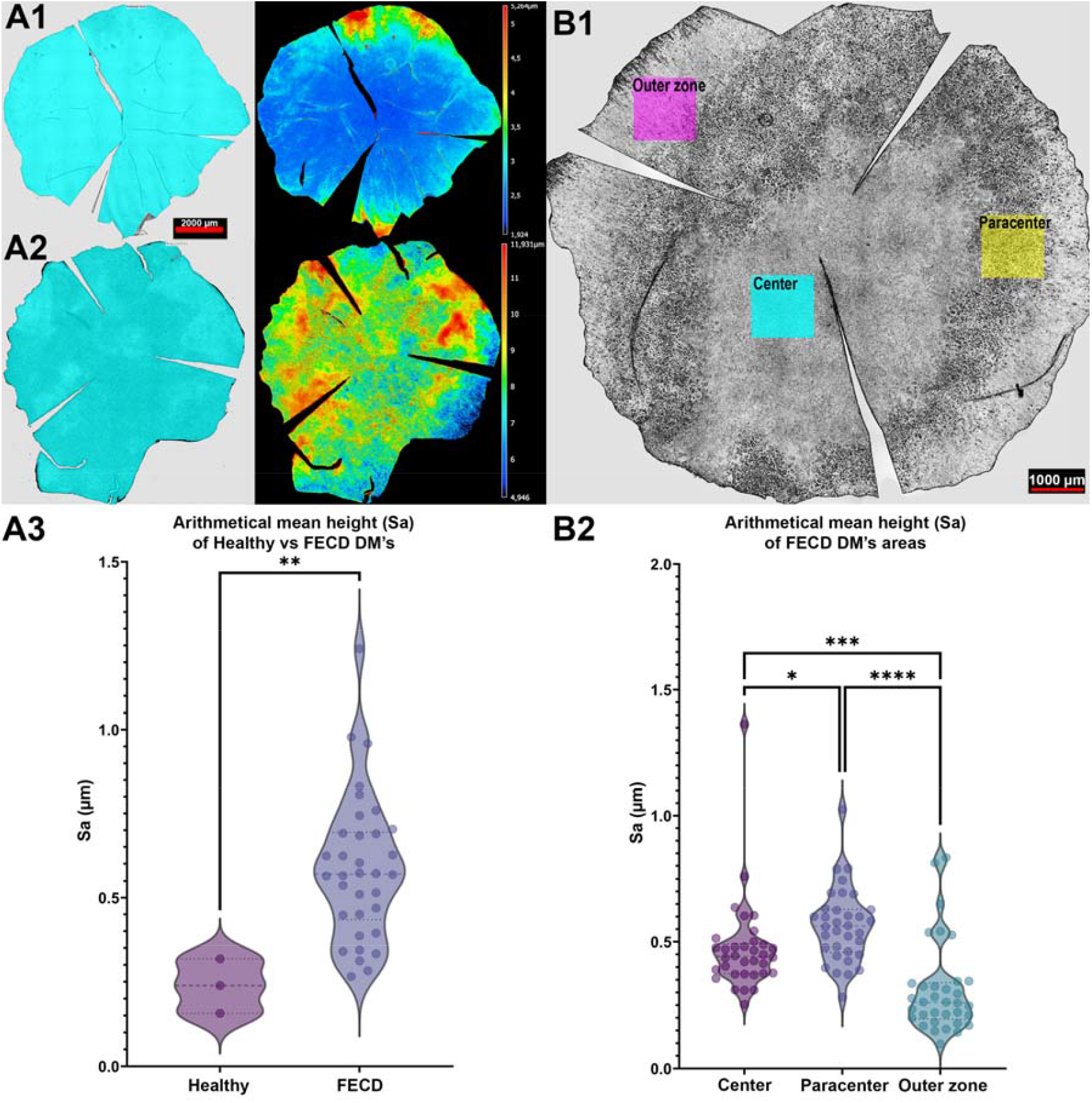
Whole-membrane and regional surface roughness analysis of healthy and FECD Descemet’s membranes (DM). **(A1–A2):** Representative whole-DM images from a healthy donor (A1) and from an FECD patient (A2). Roughness was measured over the entire highlighted area and displayed as elevation maps (10x white-light interferometry, scale bars: A1, A2 = 2000 µm, B1 = 1000 µm). **(A3):** Violin plot of areal arithmetical mean height (Sa) for healthy (n = 3) and FECD (n = 34) whole DMs (P-value = 0.0018), unpaired t-test with welch’s correction. **(B1):** Representative brightfield image of an FECD DM showing the three regions used for regional roughness analysis: center (blue), paracenter (yellow), and outer zone (magenta) (scale bar: 1000 µm). **(B2):** Violin plot of Sa in the three regions across the 34 FECD DM (P-value = 0.0423 [*], 0.0007 [***] and < 0.0001 [****]), Kruskal-Wallis ANOVA with Dunn’s multiple comparison test.

**Figure 3.**
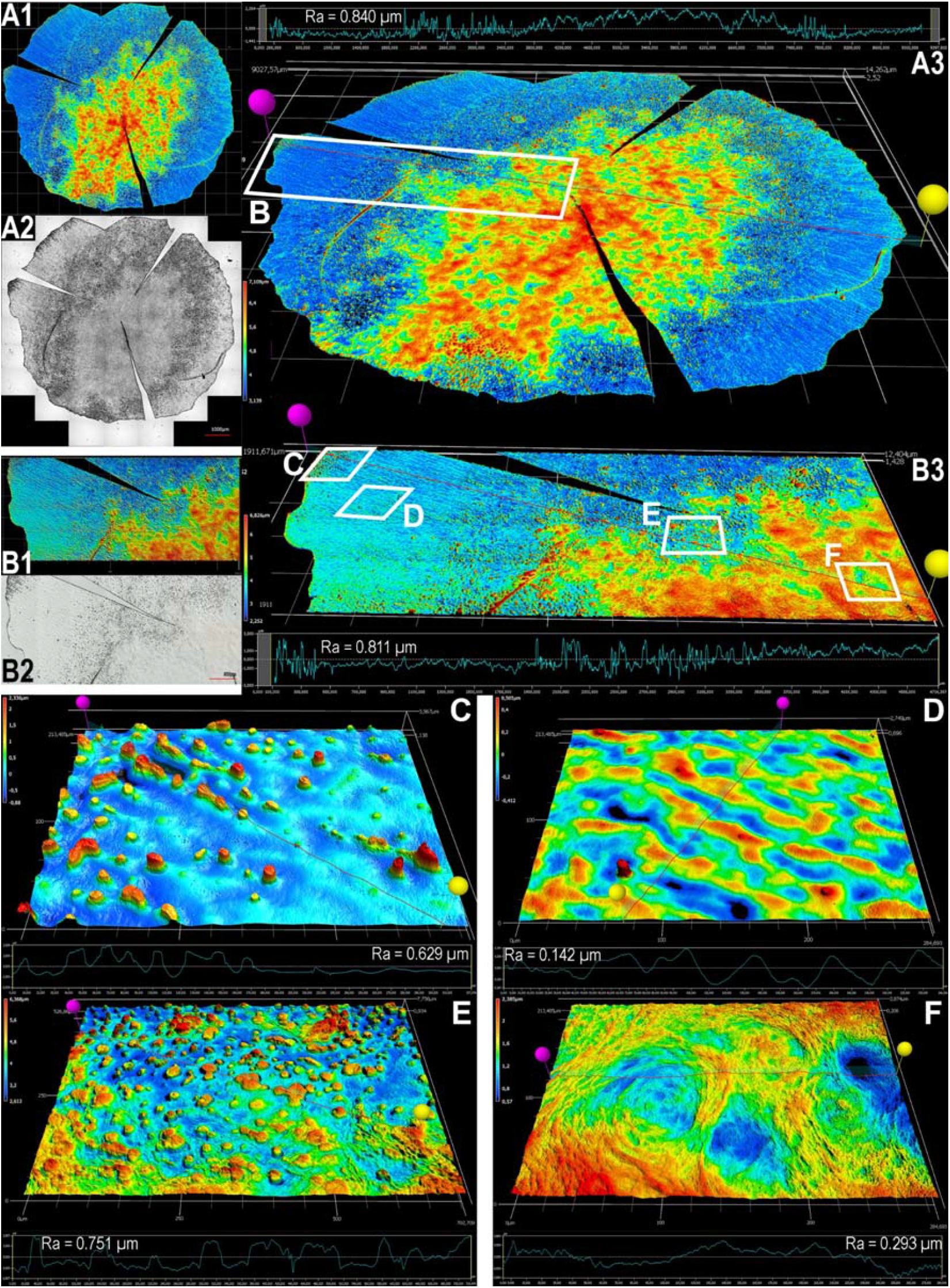
Detailed multi-scale nano-topographic analysis of a decellularized Fuchs Endothelial Corneal Dystrophy Descemet’s membrane (DM). **(A):** Whole DM reconstructed by stitching ×10 white-light interferometry images (9 × 5 tiles), scale bar: 1000 µm. **(B):** Zoomed region corresponding to panel B in A3, reconstructed by stitching ×20 confocal scans (7 × 4 tiles), scale bar: 500 µm. For A and B, **(1):** 3D elevation map of the scanned area; **(2):** brightfield image; **(3):** 3D rendering with a linear height profile along the section between the pink and yellow pins (red line). Ra: linear arithmetic mean height of the profile. **(C–E):** Zoomed regions (C, D, E in B3) (×50 laser scanning). **(F):** Zoomed region (F in B3) acquired at (×20 laser scanning).

**Figure 4.**
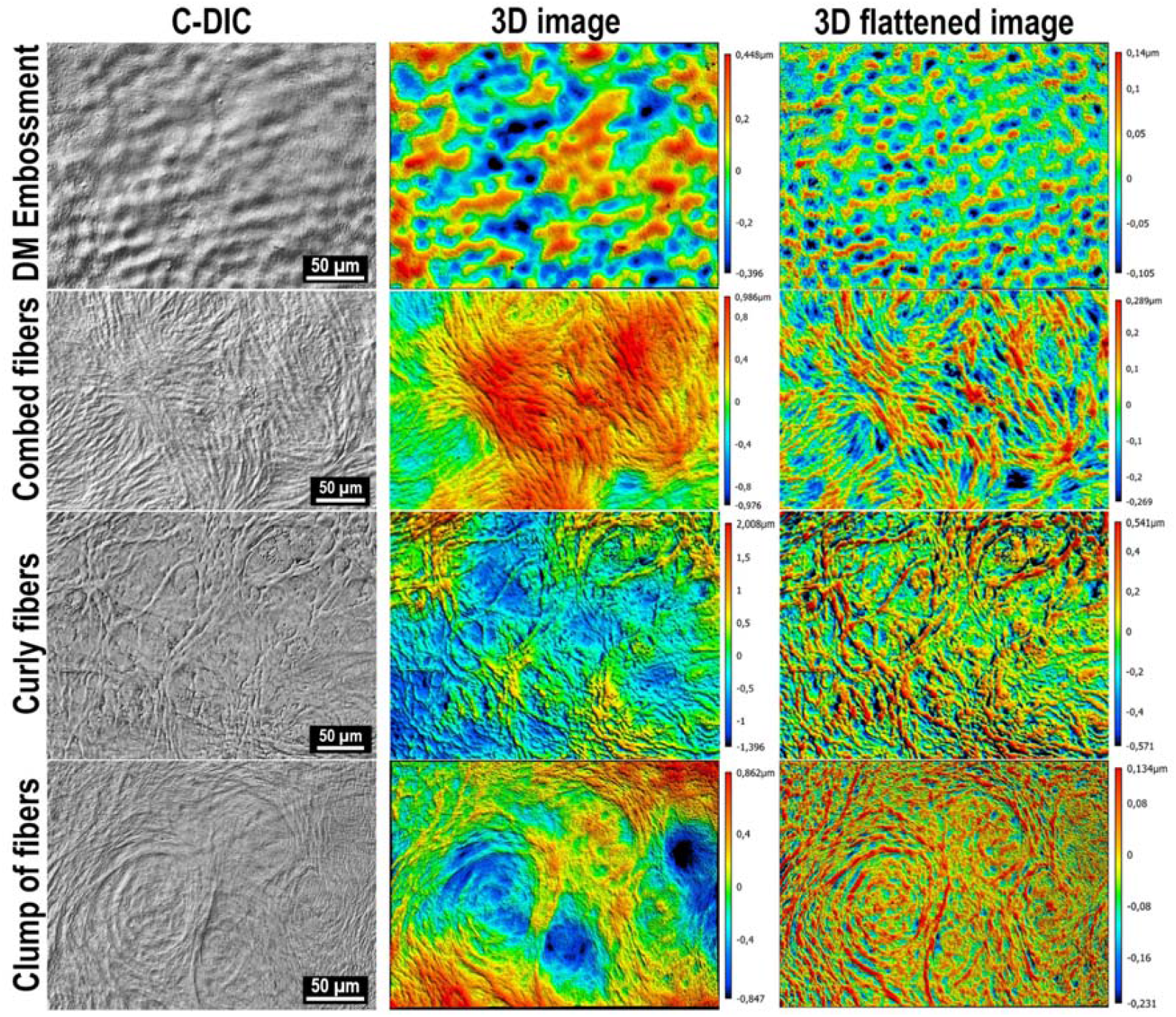
Matrix-related surface structures of Fuchs Endothelial Corneal Dystrophy Descemet’s membrane (×50 laser scanning confocal microscopy). C-DIC images were generated from brightfield images using the KEYENCE software. 3D images were reconstructed from surface analysis after reference-plane levelling. Flattened 3D images were reconstructed after wavefront removal to highlight local topography.

Roughness measurements were performed either on the whole DM or on localised regions of interest (1 µm^2^) focused on selected lesions within three regions called: center, paracenter, and outer. In most selected FECD cases, the central region was characterised by guttae covered by an additional matrix layer, the posterior fibrillar layer (PFL), which obscured guttae details. The paracentral region was immediately adjacent, with large-diameter guttae not covered by PFL. The outer region extended towards the sample edge and lacked confluent guttae and PFL. These regions were not defined by fixed distances from the sample center because their location and extent varied depending on Descemetorhexis centration and disease severity/phenotype. In rare cases (4 specimens), there was no PFL, but only isolated guttae, the centre of which was identified where the lesion load was greatest.

Surface roughness was quantified using the surface arithmetical mean height (Sa) (µm), as defined by ISO 25178: the arithmetic mean of the absolute height deviations from the mean plane. Higher (Sa) values indicate a rougher surface. For line-based (2D) profilometry, the roughness average (Ra) was used (ISO 21920), corresponding to the arithmetic mean height along a line profile.

### Statistical analysis

Whole-DM roughness comparisons between FECD and controls were performed using a non-parametric Mann–Whitney test. Comparisons across the three FECD regions (center, paracenter, outer) were performed using Kruskal–Wallis test with Dunn’s post hoc correction for multiple comparisons. Data analysis and graphs were generated using GraphPad Prism v10.6.1 (La Jolla, CA, USA).

## RESULTS

### Whole-DM topographic mapping by white-light interferometry

White-light interferometry using a ×10 objective enabled imaging of entire DMs by stitching adjacent tiles. Acquisition time was approximately 90 minutes per 8–10 mm diameter specimen. This approach allowed visualisation of guttae height, generation of elevation maps, and computation of roughness parameters over the whole DM. Roughness could be quantified from a line profile (Ra), over a defined surface or the entire specimen (Sa).

Fig. 1 showed brightfield images, elevation maps, and representative height profiles from whole DMs obtained from healthy donors without and with CECs (Fig. 1A and 1B, respectively), as well as from six FECD patients illustrating the diversity of topographic phenotypes (Fig. 1C–H).

### Surface roughness analysis

Whole-DM roughness analysis was performed on 34 FECD samples and 3 healthy samples (Fig. 2A). The analysed surface area was 61 ± 11 mm^2^. Samples did not exhibit mounting-related folds likely to bias measurements. Mean age did not differ significantly between groups (p > 0.05).

Median surface arithmetical mean height (Sa, median ± IQR) was significantly higher in FECD (Sa = 0.571 ± 0.259 µm) than in controls (Sa = 0.239 ± 0.161 µm) (p = 0.0018) (Height maps from all analysed DMs are provided in Supplementary 5).

Within a given specimen, roughness differed across the central, paracentral, and outer regions (Fig. 2B). Arithmetical mean height differed significantly between the central and paracentral regions (p = 0.0423), between the central and outer zone (p = 0.0007) and between the paracentral and outer regions (p < 0.0001). Values (Median ± IQR) were: central Sa = 0.442 ± 0.112 µm ; paracentral Sa = 0.562 ± 0.170 µm ; outer zone Sa = 0.261 ± 0.143 µm (Region delineations for all samples are shown in Supplementary 6).

### High-magnification confocal imaging of specific structures

3D confocal microscopy revealed multiple types of pathological elementary structures within the same FECD DM at higher magnification (×20, ×50) (Fig. 3). Roughness analyses were performed from line profiles and/or areal measurements at each magnification.

### Peripheral Striae

These consisted of linearly organised, radially oriented protrusions located at the sample edge. They were interrupted by the surgical tear, indicating that they likely extended further into the corneal periphery. They formed ridges several hundred microns in length, with height decreasing towards the centre (Fig. 3C).

### Furrows / Embossments

Furrows and embossments (Fig. 3D, Fig. 4) were observed in guttae-free regions, most often in the outer sample area and/or in the interval between the peripheral striae and the central confluent guttae zone. They appeared either as a bumpy DM surface with a succession of small, relatively well-delineated depressions (embossments), as shallow ridges/grooves with a radial distribution (furrows), or as a combination of both. Along furrow edges -and increasingly towards the center-isolated guttae appeared, radially aligned, with progressively increasing diameter and decreasing spacing.

### Fiber structures

In some certain FECD forms (presumed from more severely affected patients), an additional abnormal matrix deposit was present between and over guttae, forming the posterior fibrillar layer (PFL) in the centre of the cornea. 3D confocal microscopy enabled characterisation of the arrangement and dimensions of the fine, dense fibrillar structures within the PFL (Fig. 3E). It was possible to overcome the relief of the underlying guttae by applying localised digital flattening in order to isolate the excess thickness of the PFL and its patterns in a single plan (“wavefront removal”; Supplementary Data S2). Several forms were observed (Fig. 4), including curly fibers, combed fibers, and clump of fibers.

### Guttae

Multiple guttae morphologies were observed, co-existing within a given DM or present only in certain patients (Fig. 3C, 3E): isolated guttae; aligned guttae; donut-like guttae with central depression (suggesting a central collapse or remodeling); coalescent guttae and fusing guttae sometimes connected by inter-guttae bridges (Fig. 5).

**Figure 5.**
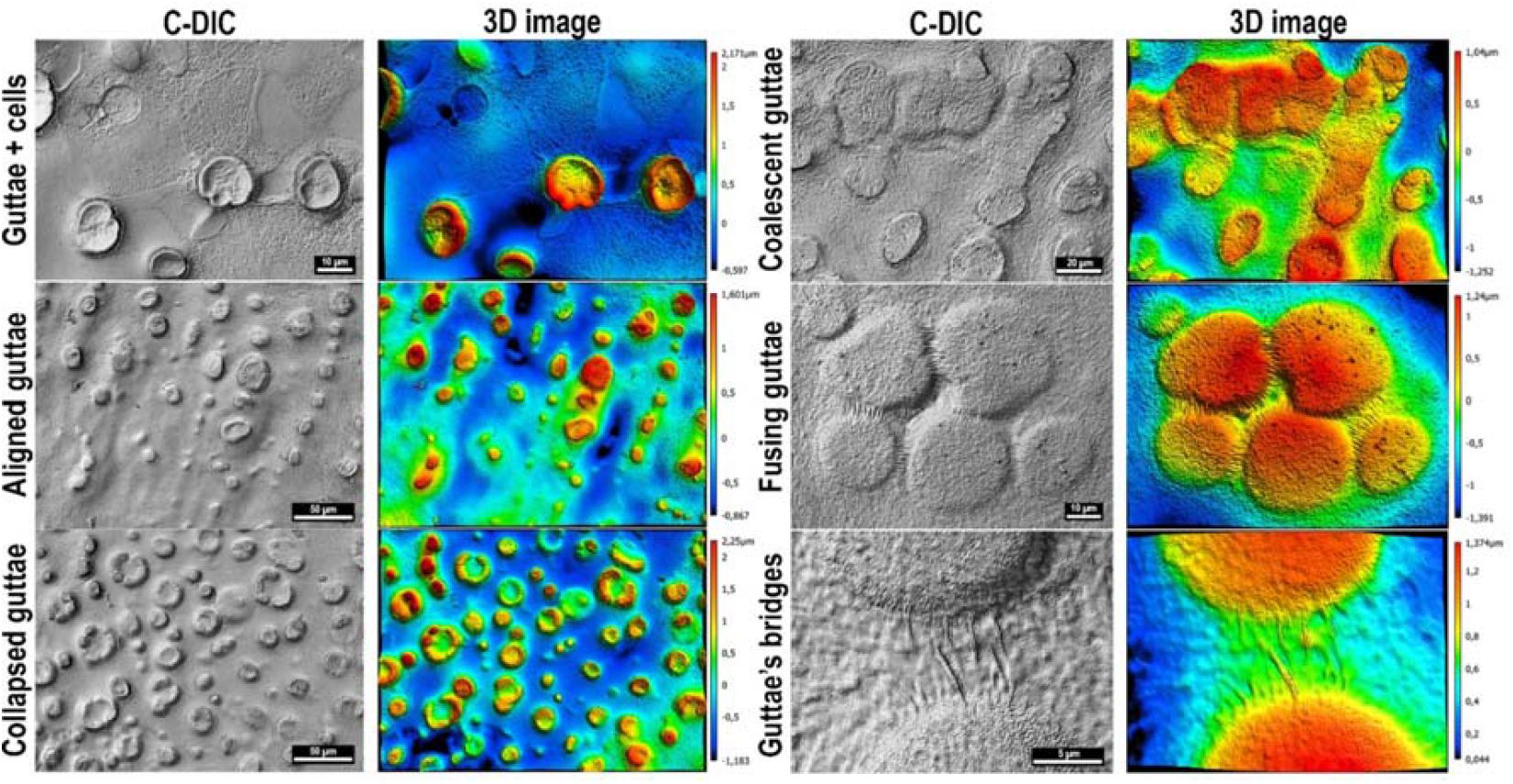
Guttae-associated structures of Fuchs Endothelial Corneal Dystrophy Descemet’s membrane (×20, ×50, and ×150 laser scanning confocal imaging). C-DIC images were generated from brightfield images using the KEYENCE software. 3D images were reconstructed from surface analysis after reference-plane levelling.

## DISCUSSION

To the best of our knowledge, this study is the first to image DM relief on whole specimens of 8 to 10 mm in diameter, corresponding approximately to half of the total Descemet surface. This high-resolution 3D topography over an extended surface was enabled by two complementary optical modalities integrated within a single instrument.

First, white-light interferometry at ×10 magnification provides wide-field imaging suitable for large-area acquisition through automated tile stitching. Despite the low magnification, optical coherence enables nanometre axial resolution and micrometre lateral resolution, allowing quantification of relief variations relevant to FECD histopathology, as axial resolution mainly depends on wavelength whereas lateral resolution depends on the optical system [19].

Second, the integration of laser scanning confocal optics (×20 to ×150) complements these observations. Although the field of view is smaller, confocal section selectivity improves lateral resolution and enables targeted imaging of regions of interest previously identified by interferometry. Together, interferometry provides highly resolved elevation mapping over large areas, while confocal imaging provides high lateral resolution in targeted regions, matched to the size of histological structures. Their combination supports a particularly relevant multiscale approach for FECD.

This methodology addresses an unmet need in current imaging. Transmission electron microscopy has described the ultrastructure of pathological DM layers and guttae for decades [6], but requires lengthy preparation and provides extremely limited sampling (on the order of tens of µm^2^), most often in transverse sections. Conversely, specular microscopy and *in vivo* confocal microscopy are fast, analyse areas of only a few hundred µm^2^ and do not provide reliable quantitative 3D topography measurements of the DM.

Beyond visualisation, our approach enables extraction of quantitative roughness parameters across scales. It is consistent with recent ex vivo work using chromatic confocal microscopy to quantify guttae topography [20], which reported comparable axial and lateral resolution but was limited to 200 µm × 200 µm acquisition windows without the ability to survey larger areas via multi-magnification imaging or tile stitching. Another strength of the present approach is post-processing capability, which can correct sample-tilt artefacts and, importantly, isolate relief patterns associated with the PFL by reducing the influence of the higher-amplitude guttae topography (“wavefront removal”).

These two microscopy modalities enable characterisation of the full spectrum of DM modifications that we recently described in a series of 500 FECD specimens using transmitted brightfield microscopy[8]. In this subset (34/500), we again observe distinct topographic signatures across patients, supporting the potential of large-scale quantitative topography to refine histological subgrouping. Precise subgroup characterisation is expected to impact both basic research (improved understanding of pathophysiology; identification of subgroup-specific mechanisms) and clinical care (personalised monitoring; selection of homogeneous populations for clinical trials).

We also focused on three structures that we recently described and for which we are formulating new pathophysiological hypotheses: peripheral striae, DM furrows/embossments, and PFL fibres structures [8], [21]. The radial organisation of striae and furrows/embossments mirrors the physiological architecture of healthy endothelium, which shows radial columns of CECs between Hassall–Henle bodies in the far periphery [22]. We previously suggested that cell migration may play a key role in FECD pathophysiology, in addition to mechanisms such as increased oxidative stress [23], mitochondrial dysfunction [24], and endoplasmic reticulum stress/unfolded protein response [25]. An abnormal CEC population with exaggerated matrix secretion (and/or abnormal remodelling capacity) could progressively migrate towards the centre and generate these radial structures. The progressive concentration of abnormalities towards the corneal centre could reflect gradual migrational slowing as cells accumulate centrally. Alternatively, all CECs may retain a centripetal radial organisation from embryonic migration that is not apparent in healthy endothelium. Preferential FECD expression in specific CEC subpopulations could then reveal this primordial organisation. These two hypotheses are not mutually exclusive and may be self-reinforcing.

The presence of a central PFL is described as a marker of FECD severity, as it is associated with severe CEC dysfunction [26], [27]. It preferentially appears in regions with large, confluent guttae, which have been associated in vitro with substantial cellular stress and reduced endothelial viability [28]. Here, we show that roughness in this central region -where guttae are buried under PFL-is significantly lower than in the adjacent paracentral region, where roughness peaks due to thick guttae not covered by PFL. This relatively smooth central surface may have more limited optical consequences than a rough surface and could help explain why some patients maintain acceptable visual acuity for years, before chronic oedema develops as CEC function collapses.

Overall, the innovative approach presented here complements conventional histological observations by providing a three-dimensional, quantitative assessment of the many matrix abnormalities characteristic of FECD. High-resolution analysis across the entire Descemetic surface should help generate new pathophysiological hypotheses and identify patient subgroups with distinct disease trajectories or differential responses to emerging therapies.

## Data Availability

All data produced in the present study are available upon reasonable request to the authors

## References

1. Van Meter W, Buckman N, Philippy B, DeMatteo J, America on behalf of the EBA of. 2024 Eye Banking Statistical Report—Executive Summary. Eye Banking and Corneal Transplantation. juin 2025;4(2):e0042. doi:10.1097/ebct.0000000000000042

2. Ong Tone S, Kocaba V, Böhm M, Wylegala A, White TL, Jurkunas UV. Fuchs endothelial corneal dystrophy: The vicious cycle of Fuchs pathogenesis. Prog Retin Eye Res. janv 2021;80:100863. doi:10.1016/j.preteyeres.2020.100863 PubMed PMID: 32438095; PubMed Central PMCID: PMC7648733.

3. Aiello F, Gallo Afflitto G, Ceccarelli F, Cesareo M, Nucci C. Global Prevalence of Fuchs Endothelial Corneal Dystrophy (FECD) in Adult Population: A Systematic Review and Meta-Analysis. J Ophthalmol. 2022;2022:3091695. doi:10.1155/2022/3091695 PubMed PMID: 35462618; PubMed Central PMCID: PMC9023201.

4. Barrera-Sanchez M, Hernandez-Camarena JC, Ruiz-Lozano RE, Valdez-Garcia JE, Rodriguez-Garcia A. Demographic profile and clinical course of Fuchs endothelial corneal dystrophy in Mexican patients. Int Ophthalmol. 1 avr 2022;42(4):1299□309. doi:10.1007/s10792-021-02117-0

5. Kocaba V, Katikireddy KR, Gipson I, Price MO, Price FW, Jurkunas UV. Association of the Gutta-Induced Microenvironment With Corneal Endothelial Cell Behavior and Demise in Fuchs Endothelial Corneal Dystrophy. JAMA Ophthalmol. 1 août 2018;136(8):886□92. doi:10.1001/jamaophthalmol.2018.2031 PubMed PMID: 29852040; PubMed Central PMCID: PMC6142944.

6. Bourne WM. The Ultrastructure of Descemet’s Membrane: III. Fuchs’ Dystrophy. Arch Ophthalmol. 1 déc 1982;100(12):1952. doi:10.1001/archopht.1982.01030040932013

7. Xia D, Zhang S, Nielsen E, Ivarsen AR, Liang C, Li Q, Thomsen K, Hjortdal JØ, Dong M. The Ultrastructures and Mechanical Properties of the Descement’s Membrane in Fuchs Endothelial Corneal Dystrophy. Sci Rep. 16 mars 2016;6(1):23096. doi:10.1038/srep23096

8. Vaitinadapoulé H, Onitiu D, Maurin C, Travers G, Crouzet E, Dorado-Cortez O, Poinard S, He Z, Forest F, Ollier E, Touraine R, Gain P, Perone JM, Thuret G, Group (FFSG) the FFS. Pathological classification of Fuchs endothelial corneal dystrophy in several types and their relationships with CTG18.1 expansion repeats [Internet]. medRxiv; 2025 [cité 23 nov 2025]. p. 2025.07.14.25330988. Disponible sur: https://www.medrxiv.org/content/10.1101/2025.07.14.25330988v1 doi:10.1101/2025.07.14.25330988

9. Maeno S, Lewis PN, Young RD, Oie Y, Nishida K, Quantock AJ. Imaging pathology in archived cornea with Fuchs’ endothelial corneal dystrophy including tissue reprocessing for volume electron microscopy. Sci Rep. 30 déc 2024;14(1):31786. doi:10.1038/s41598-024-82888-5 PubMed PMID: 39738318; PubMed Central PMCID: PMC11685999.

10. Yuen HKL, Rassier CE, Jardeleza MSR, Green WR, de la Cruz Z, Stark WJ, Gottsch JD. A Morphologic Study of Fuchs Dystrophy and Bullous Keratopathy. Cornea. avr 2005;24(3):319. doi:10.1097/01.ico.0000148288.53323.b2

11. Laing RA, Leibowitz HM, Oak SS, Chang R, Berrospi AR, Theodore J. Endothelial mosaic in Fuchs’ dystrophy. A qualitative evaluation with the specular microscope. Arch Ophthalmol. janv 1981;99(1):80□3. doi:10.1001/archopht.1981.03930010082007 PubMed PMID: 6970032.

12. Fujimoto H, Maeda N, Soma T, Oie Y, Koh S, Tsujikawa M, Nishida K. Quantitative regional differences in corneal endothelial abnormalities in the central and peripheral zones in Fuchs’ endothelial corneal dystrophy. Invest Ophthalmol Vis Sci. 24 juill 2014;55(8):5090□8. doi:10.1167/iovs.14-14249 PubMed PMID: 25061116.

13. Mustonen RK, McDonald MB, Srivannaboon S, Tan AL, Doubrava MW, Kim CK. In vivo confocal microscopy of Fuchs’ endothelial dystrophy. Cornea. sept 1998;17(5):493□503. doi:10.1097/00003226-199809000-00006 PubMed PMID: 9756443.

14. Beierlein G, Haas L, Hahnel S, Schmidt M, Rosentritt M. Influence of cement type, excess removal, and polishing on the cement joint. Quintessence Int. 28 févr 2024;55(2):98□105. doi:10.3290/j.qi.b4780239 PubMed PMID: 38108419.

15. Morán MC, Cirisano F, Ferrari M. 3D profilometry and cell viability studies for drug response screening. Materials Science and Engineering: C. 1 oct 2020;115:111142. doi:10.1016/j.msec.2020.111142

16. ISO 25178-2:2021(en), Geometrical product specifications (GPS) — Surface texture: Areal — Part 2: Terms, definitions and surface texture parameters [Internet]. [cité 3 avr 2026]. Disponible sur: https://www.iso.org/obp/ui/#iso:std:iso:25178:-2:ed-2:v1:en

17. Borderie VM, Sabolic V, Touzeau O, Scheer S, Carvajal-Gonzalez S, Laroche L. Screening human donor corneas during organ culture for the presence of guttae. Br J Ophthalmol. mars 2001;85(3):272□6. doi:10.1136/bjo.85.3.272 PubMed PMID: 11222329; PubMed Central PMCID: PMC1723890.

18. Dapena I, Moutsouris K, Droutsas K, Ham L, van Dijk K, Melles GRJ. Standardized « no-touch » technique for descemet membrane endothelial keratoplasty. Arch Ophthalmol. janv 2011;129(1):88□94. doi:10.1001/archophthalmol.2010.334 PubMed PMID: 21220634.

19. Fujimoto JG, Pitris C, Boppart SA, Brezinski ME. Optical coherence tomography: an emerging technology for biomedical imaging and optical biopsy. Neoplasia. 2000;2(1□2):9□25. doi:10.1038/sj.neo.7900071 PubMed PMID: 10933065; PubMed Central PMCID: PMC1531864.

20. Vaïtinadapoulé H, Poinard S, He Z, Pascale Hamri A, Thomas J, Gain P, Thuret JY, Mascarelli F, Thuret G. Nanotopography by chromatic confocal microscopy of the endothelium in Fuchs endothelial corneal dystrophy, pseudophakic bullous keratopathy and healthy corneas. British Journal of Ophthalmology. 15 sept 2023;108:bjo-2023. doi:10.1136/bjo-2023-323297

21. Thuret G, Ain A, Koizumi N, Okumura N, Gain P, He Z. Radial Endothelial Striae Over 360 Degrees in Fuchs Corneal Endothelial Dystrophy: New Pathophysiological Findings. Cornea. 1 déc 2021;40(12):1604□6. doi:10.1097/ICO.0000000000002666 PubMed PMID: 33591033.

22. He Z, Campolmi N, Gain P, Ha Thi BM, Dumollard JM, Duband S, Peoc’h M, Piselli S, Garraud O, Thuret G. Revisited microanatomy of the corneal endothelial periphery: new evidence for continuous centripetal migration of endothelial cells in humans. Stem Cells. nov 2012;30(11):2523□34. doi:10.1002/stem.1212 PubMed PMID: 22949402.

23. Jurkunas UV, Bitar MS, Funaki T, Azizi B. Evidence of oxidative stress in the pathogenesis of fuchs endothelial corneal dystrophy. Am J Pathol. nov 2010;177(5):2278□89. doi:10.2353/ajpath.2010.100279 PubMed PMID: 20847286; PubMed Central PMCID: PMC2966787.

24. Miyai T. Fuchs Endothelial Corneal Dystrophy and Mitochondria. Cornea. nov 2018;37 Suppl 1:S74□7. doi:10.1097/ICO.0000000000001746 PubMed PMID: 30252683.

25. Engler C, Kelliher C, Spitze AR, Speck CL, Eberhart CG, Jun AS. Unfolded protein response in fuchs endothelial corneal dystrophy: a unifying pathogenic pathway? Am J Ophthalmol. févr 2010;149(2):194–202.e2. doi:10.1016/j.ajo.2009.09.009 PubMed PMID: 20103053; PubMed Central PMCID: PMC2813215.

26. Hribek A, Clahsen T, Horstmann J, Siebelmann S, Loreck N, Heindl LM, Bachmann BO, Cursiefen C, Matthaei M. Fibrillar Layer as a Marker for Areas of Pronounced Corneal Endothelial Cell Loss in Advanced Fuchs Endothelial Corneal Dystrophy. Am J Ophthalmol. févr 2021;222:292□301. doi:10.1016/j.ajo.2020.09.030 PubMed PMID: 32971030.

27. Özer O, Mestanoglu M, Howaldt A, Clahsen T, Schiller P, Siebelmann S, Reinking N, Cursiefen C, Bachmann B, Matthaei M. Correlation of Clinical Fibrillar Layer Detection and Corneal Thickness in Advanced Fuchs Endothelial Corneal Dystrophy. J Clin Med. 17 mai 2022;11(10):2815. doi:10.3390/jcm11102815 PubMed PMID: 35628952; PubMed Central PMCID: PMC9144691.

28. Kocaba V, Katikireddy KR, Gipson I, Price MO, Price FW, Jurkunas UV. Association of the Gutta-Induced Microenvironment With Corneal Endothelial Cell Behavior and Demise in Fuchs Endothelial Corneal Dystrophy. JAMA Ophthalmol. août 2018;136(8):886□92. doi:10.1001/jamaophthalmol.2018.2031 PubMed PMID: 29852040; PubMed Central PMCID: PMC6142944.

