## Supplementary 1 for "Three-dimensional topography of Descemet’s membrane in Fuchs endothelial corneal dystrophy using laser scanning confocal microscopy and white-light interferometry"

**Supplementary 1: The French Fuchs Study Group. Twenty-five ophthalmology departments from the following hospitals (listed in alphabetical order by hospital and including surgeons):**

| Surgeons | Hospital, city | Country |
|---|---|---|
| Dr Diane Bernheim<br>Prof. Christophe Chiquet | University Hospital of Grenoble, Grenoble | France |
| Prof. Vincent Borderie | Hôpital National des 15-20, IHU ForeSight, GRC 32, Transplantation et Thérapies Innovantes de la Cornée, TTIC, INSERM-DGOS CIC 1423, Paris | France |
| Prof. Tristan Bourcier | University Hospital, Nouvel Hôpital Civil, Strasbourg | France |
| Prof. Jean-Louis Bourges | University Hospital Cochin, Paris | France |
| Prof. Frédéric Chiambaretta | University Hospital Gabriel Montpied, Clermont-Ferrand | France |
| Prof. Béatrice Cochener | University Hospital Morvan, Brest | France |
| Prof. Louis Arnould<br>Dr Florian Baudin<br>Prof. Catherine Creuzot | University Hospital of Dijon | France |
| Prof. Vincent Daien | University Hospital of Montpellier | France |
| Prof. Alexandre Denoyer | University Hospital Robert Debré, Reims | France |
| Prof. Bernard Duchesne | University Hospital Sart Tilman, Liège | Belgium |
| Dr Nicolas Duquesne | Kleber Ophthalmology Centre, Lyon | France |
| Prof. Pierre Fournie | University Hospital Purpan, Toulouse | France |
| Prof. Anne-Sophie Gauthier | University Hospital of Besançon | France |
| Prof. Philippe Gain<br>Prof. Gilles Thuret | University Hospital of Saint-Etienne | France |
| Prof. Louis Hoffart | Clinique Monticelli-Vélodrome, Marseille | France |
| Dr François Majo | Ophthalmology Centre, Lausanne | Switzerland |
| Prof. Marc Muraine | University Hospital Charles Nicolle, Rouen | France |
| Dr Romain Mouchel | Kleber Ophthalmology Centre, Lyon | France |
| Dr Jean Marc Perone | Hospital Center Metz-Thionville | France |
| Prof. Jean Claude Quintyn | University Hospital Caen/Normandie | France |
| Dr Alexandra Rabot | Hospital Center, Antibes Juan les Pins | France |
| Dr Alain Saad<br>Dr Damien Gatinel | Rothschild Foundation Hospital, Paris | France |
| Dr Pierre-Yves Santiago<br>Dr Jean-Michel Bosc | Institut ophtalmologique de l'ouest, Clinique Jules Verne, Nantes | France |
| Prof. David Toubould | University Hospital Pellegrin, Bordeaux | France |
| Dr Bertrand Vabres | University Hospital Hôtel Dieu, Nantes | France |
