## Supplementary 2 for "Three-dimensional topography of Descemet’s membrane in Fuchs endothelial corneal dystrophy using laser scanning confocal microscopy and white-light interferometry"

**Supplementary 2:** List of Descemet's membrane specimens analysed, including diagnosis (healthy/FECD), fixation, associated figure(s), and histological type (from [8]).

| ID | Healthy/ FECD | Fixation | Figure in article | Histological type previously described |
|---|---|---|---|---|
| F89 | FECD | PFA | Fig2 A3, B2, B3 | Radial |
| F179 | FECD | PFA | Fig2 A3, B2, B3 | Guttae only |
| F209 | FECD | PFA | Fig2 A3, B2, B3 | Center |
| F255 | FECD | BSS | Fig2 A3, B2, B3 | Fused |
| F274 | FECD | PFA | Fig2 A3, B2, B3 | Epicenter |
| F275 | FECD | PFA | Fig2 A2, A3, B2, B3 | Epicenter |
| F383 | FECD | WATER | Fig2 A3, B2, B3 | Center |
| F387 | FECD | BSS | Fig2 A3, B2, B3 | Radial |
| F588 | FECD | WATER | Fig2 A3, B2, B3; Fig4 Curly fibers | Radial |
| F606 | FECD | WATER | Fig2 A3, B2, B3 | Radial |
| F663 | FECD | BSS | Fig1 E; Fig2 A3, B2, B3 | Radial |
| F669 | FECD | BSS | Fig2 A3, B2, B3 | Outlier |
| F671 | Healthy | WATER | Fig2 A1, A3 | Healthy |
| F673 | Healthy | WATER | Fig2 A3 | Healthy |
| F692 | FECD | PFA | Fig2 A3, B2, B3; Fig5 Fusing guttae, Guttae's bridges | Healthy |
| F700 | Healthy | PFA | Fig1 B (DM covered by cells) | Healthy |
| F710 | FECD | CMX | Fig2 A3, B2, B3 | Outlier |
| F750 | FECD | WATER | Fig1 F; Fig2 A3, B2, B3 | Outlier |
| F768 | Healthy | WATER | Fig1 A | Healthy |
| F788 | FECD | WATER | Fig1 D; Fig2 A3, B2, B3 | Radial |
| F795 | FECD | WATER | Fig2 A3, B2, B3 | Radial |
| F823 | FECD | WATER | Fig2 A3, B2, B3 | Radial |
| F828 | FECD | WATER | Fig2 A3, B2, B3 | Epicenter |
| F860 | FECD | WATER | Fig5 Fused guttae | Center |
| F893 | FECD | WATER | Fig2 A3, B1, B2, B3; Fig3; Fig4 Clump of fibers | Center |
| F899 | FECD | CMX | Fig2 A3, B2, B3 | Center |
| F945 | FECD | WATER | Fig2 A3, B2, B3 | Epicenter |

| | | | | |
|---|---|---|---|---|
| F1074 | FECD | BSS | Fig1 C; Fig4 Combed fiber | Radial |
| F1104 | FECD | BSS | Fig2 A3, B2, B3 | Guttae only |
| F1116_2 | FECD | BSS | Fig2 A3, B2, B3 | Outlier |
| F1158 | FECD | WATER | Fig5 Aligned guttae, Collapsed guttae | Epicenter |
| F1196 | FECD | BSS | Fig2 A3, B2, B3; Fig4 DM Embossment | Guttae only |
| F1199 | FECD | BSS | Fig2 A3, B2, B3 | Guttae only |
| F1201 | FECD | BSS | Fig2 A3, B2, B3 | Center / Radial |
| F1345 | FECD | WATER | Fig1 H; Fig2 A3, B2, B3 | Radial / Epicenter |
| F1366 | FECD | WATER | Fig1 G; Fig2 A3, B2, B3 | Radial / Guttae only / Fused |
| F1376 | FECD | WATER | Fig2 A3, B2, B3 | Outlier |
| F1378 | FECD | WATER | Fig2 A3, B2, B3 | Center |
| F1379 | FECD | WATER | Fig2 A3, B2, B3 | Radial |
| F1391 | FECD | WATER | Fig2 A3, B2, B3 | Radial |
| F1400 | FECD | WATER | Fig2 A3, B2, B3 | Radial / Fused |
| F1434 | FECD | ALCHIMIA | Fig5 Guttae + cells | Center* |

*this specimen was not included in the series of 500 cases by Vaitinadapoule et al. (DOI: 10.1002/path.70044)
