## Supplementary 3 for "Three-dimensional topography of Descemet’s membrane in Fuchs endothelial corneal dystrophy using laser scanning confocal microscopy and white-light interferometry"

**Supplementary 3:** Objective references and resolutions by optical mode and magnification.

| | Z Resolution | XY resolution | Reference |
|---|---|---|---|
| Interferometry 10X objective | 0,01 nm | 1,318 µm | CF IC EPI Plan DI 10X WD 7,40mm NA 0,30 |
| Confocal 20X objective | 12 nm | 0,659 µm | CF IC EPI Plan 20x WD 3,10mm NA 0,46 |
| Confocal 50X objective | | 0,264 µm | CF IC EPI Plan Apo 50x WD 0,35mm NA 0,95 |
| Confocal 100X objective | | 0,132 µm | CF IC EPI Plan ELWD 100X WD 2,20mm NA 0,8 |
| Confocal 150X objective | | 0,088 µm | CF IC EPI plan Apo 150X WD 0,20mm NA 0,95 |
