## Supplementary figures and images for "Three-dimensional topography of Descemet’s membrane in Fuchs endothelial corneal dystrophy using laser scanning confocal microscopy and white-light interferometry"

### Supplementary 4

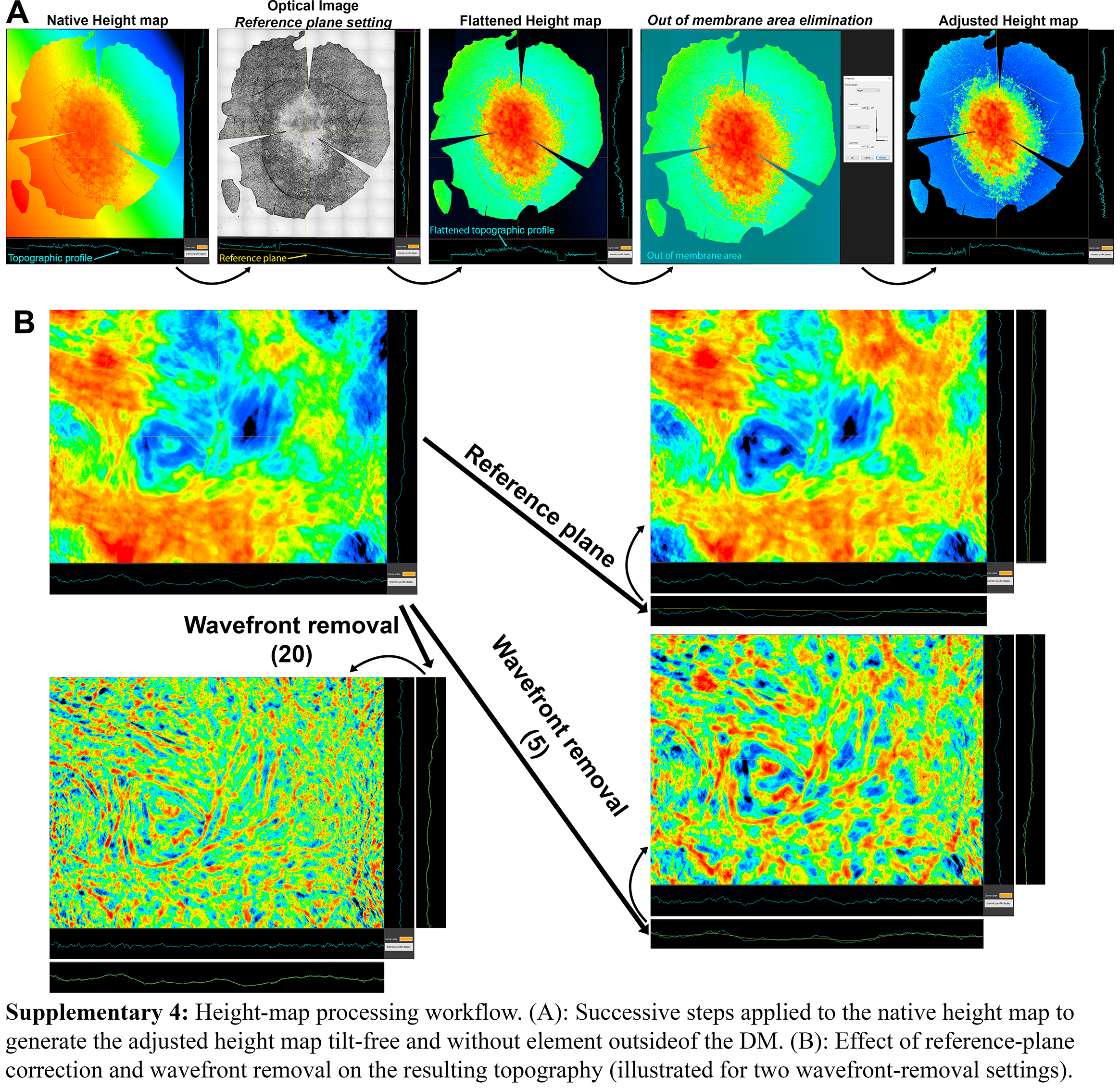

### Supplementary 5

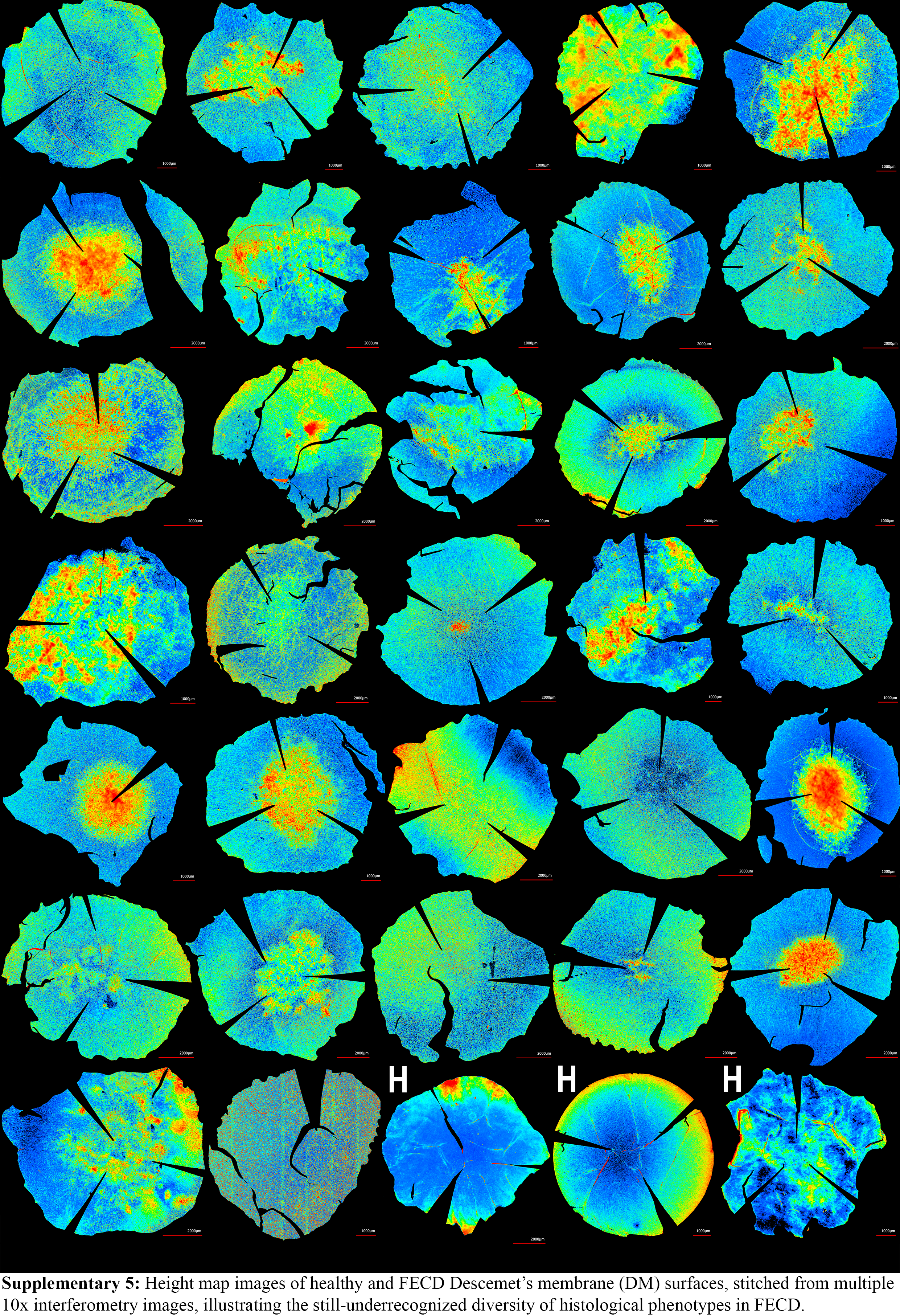

### Supplementary 6

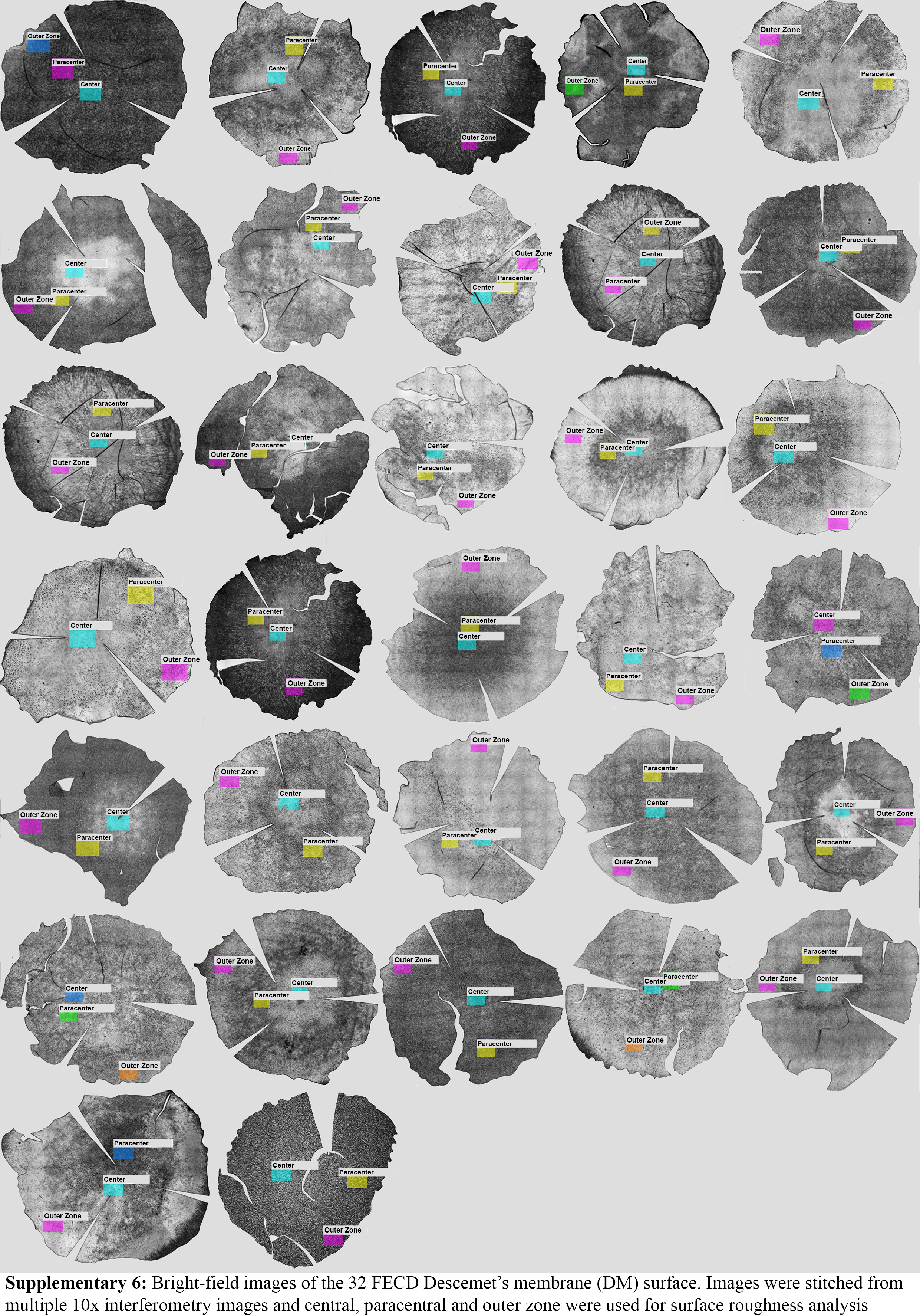
